# Impact of family support on the cognitive functions of Egyptian elderly

**DOI:** 10.1101/2023.03.02.23286696

**Authors:** Sara A. Moustafa, Nada Gaballah, Shimaa Heikal, Mohamed Salama

**Affiliations:** Institute of Global Health and Human Ecology, The American University in Cairo, Egypt; Department of Computer Science, The American University in Cairo; Faculty of Medicine, Mansoura University, Dakahlia; Atlantic Senior Fellow of Equity in Brain Health at the Global Brain Health Institute (GBHI), Trinity College Dublin, (TCD), Dublin, Ireland

**Keywords:** ageing population, cognitive functions, family support, Egyptian population, AL-SEHA

## Abstract

**Background:** Social support is essential in the daily activities of the elderly, which can impact their cognitive functions over time.

**Aim of the study:** This study investigates the hypothesis that there is a link between social support and cognitive function in the elderly living in the Egyptian community using the Arabic translation of the Survey of Health, Ageing, Retirement in Europe (SHARE) questionnaire.

**Subjects and methods:** Cross-sectional analytic study was conducted as a part of the pilot study for A Longitudinal Study of Egyptian Healthy Ageing (AL-SEHA) project. It included 299 participants (50+ years of age). Investigators collected data using the Arabic translation of the SHARE questionnaire in five Egyptian Universities. Data were then uploaded to the Social Research Centre (SRC) at the American University in Cairo (AUC).

**Results:** The prevalence of declined cognitive functions was 39.467% (95% CI: 33.89-45.04). Cognitive impairment was significantly less among university and postgraduate degree holders (p<0.001), married (p=0.0378) or contacted by their kids on a daily or weekly basis (p=0.0364).

**Conclusion:** Being married, with a university degree or higher, and increased contact frequency with children, all showed positive correlation with cognitive function in our sample. In conclusion, this research contributes to our understanding of the impact of family network and support and cognitive function in the older Egyptian population. Our findings can be a base to add on to the literature.

## 1. Introduction

Adults over 65 account for an increasing proportion of the population in developed countries (Restrepo & Rozental, 1994). This proportion is increasing and is expected to reach one in five people in developing countries by 2050 (Shetty, 2012). With this demographic shift, it is critical to investigate how to age successfully with low levels of disability, high cognitive and physical functioning, and high activity level (Depp & Jeste, 2006; Rowe & Kahn, 1997 & Urtamo et al., 2019). Cognitive functioning is an important aspect of successful <u>aging</u>, and while cognitive decline is common in older adults, the rate of change is highly variable (Fillit et al., 2002; Park, O’Connell, & Thomson, 2003; Fernández-Ballesteros & R., Benetos, 2019). Physical health (Perlmutter & Nyquist, 1990; Zelinski, Crimmins, Reynolds, & Seeman, 1998), depression (Ownby, Crocco, Acevedo, John, & Loewenstein, 2006), and physical activity (Heyn, Abreu, & Ottenbacher, 2006) are all important individual characteristics that are consistently associated with cognitive decline variability as well as, social interaction (James, Wilson, Barnes, & Bennett, 2011; Zunzunegui, 2011, Alvarado, Deindl, C., Brandt, M. and Hank, K. 2015)). However, questions remain about how these variables interact with one another and whether some, but not all, are uniquely associated with cognition over time.

However, the literature does not support the notion that living with others is beneficial to cognitive health. Some empirical studies appear to suggest that living with others may be beneficial to cognitive functioning. In line with the economic (Casey and Yamada, 2002, Bertogg, A. and Leist, A.K. 2021) and psychological benefits (De Jong Gierveld, Dykstra, and Schenk, 2012) of living with a partner in later life, some studies suggest that being in a partnership has a positive effect on cognitive functioning compared to being single (*Hkansson et al*.; *Mousavi-Nasab et al*.; *Van Gelder et al*.). Additionally, the impact of living with adult children was also studied and showed a positive influence on cognitive function although uncertain (MAZZUCO, S.T.E.F.A.N.O. et al. (2016),Zhang & Fletcher, 2021). Yet, the relationship between family status (marital status and parenthood), well-being, and psychological well-being is widely debated in academic and popular discourses (Li, M. et al. (2020). Research shows that being married or residing with a partner is associated with Improved mental health and fewer depressive symptoms in old age. (Bures et.al 2004, Gibney et.al 2017)

However, the direct pathway of impact only accounts for a portion of the effect of family relations on cognitive health. In addition to providing mental stimulation, interacting and collaborating with others can improve cognitive health by lowering psychological stress and improving mental health. (Berkman and colleagues, 2000, Arpino, B. and Bordone, V. 2014) present a conceptual model of cascading social processes in which meaningful other engagement can affect cognitive health via mediators such as psychological distress.

Although this proposed mechanism has been raised as a possible explanation for the associations between social networks and cognitive health, there has been little empirical research into it (Fratiglioni, Paillard-Borg, & Winblad, 2004, Bourassa, K.J. et al. 2015).

Moreover, contact frequency is commonly assumed to directly impact cognitive health in old age, as represented by the ‘use it or lose it’ theory. According to this viewpoint, frequent interaction with others provides ‘cognitive exercise,’ stimulating the mind and preserving cognitive functions (Hertzog, Kramer, Wilson, & Lindenberger, 2009; Hultsch, Hertzog, Small, & Dixon, 1999). Navigating social cues, organizing social gatherings and engaging in complex social discourse can all provide cognitive stimulation (Bielak, 2010; Kim & Kim, 2014). Such encounters can also improve cognitive reserve, allowing people to tolerate brain pathology without displaying obvious behavioral symptoms (Bennett, Schneider, Tang, Arnold, & Wilson, 2006).

The aspects of parenthood may thus differ depending on the life-cycle stage, implying that the positive aspects of parenthood predominate as we age. The role of children as a form of social support (Yeh, S.-C.J. and Liu, Y.-Y. 2003), among other things, may become necessary in the later stages of a person’s life (Margolis R. et.al 201, Becker, C., Kirchmaier, I. and Trautmann, S.T. 2019). But what exactly is social support? Cobb S. provides one of the most frequently cited definitions, describing social support as “information leading the subject to believe that he is cared For and adored, respected, and a member of mutual obligations.” Social support is described by the U.S. National Cancer Institute as “a network of family, friends, neighbors, and community members that are available in times of need to give psychological, physical, and financial assistance.” A social network is defined as a “set of actors and the ties that connect them” (Wesserman S. et.al, 1994), while social support describes the quantity and quality of these ties from an individual perspective. While there are numerous definitions of social support, most of them include factors such as network size and structure, the physical distance to other network members, length of relationships, frequency of contact, or function of each ties (Pearson JE et. al, 1986). According to the evidence, such social support networks are associated with less loneliness and more happiness (Litwin H. et.al, 2011, Litwin H and Stoecke KJ 2014, Şahin, D.S., Özer, Ö. And Yanardağ, M.Z. 2019))and serve as an essential buffer against stressful situations (Cohn-Schwartz, E., Levinsky, M. and Litwin, H. 2020).

A longitudinal study conducted by Bassuk and colleagues (1999) discovered that elderly people with no social ties were at a higher risk of cognitive decline than those with five or six social ties. Boult and colleagues discovered that social support was associated with a lower risk of developing disability up to 4 years later by using frequency of contact with friends and colleagues as an indicator. A prospective cohort study of 1,203 non-dementia patients was conducted. Fratiglioni and colleagues (2004) discovered that a social network reduced the incidence of dementia in people aged 75 and up for three years.

We investigate the effect of social support on cognitive function in a convenient sample of the Egyptian population. We investigated the relationship between social support and cognitive function in the elderly by controlling for individual characteristics such as age, gender, and health status.

While the results on parenthood are debatable and depend on the age of the population studied, there is widespread agreement that social support is associated with higher life satisfaction and that social networks are an important factor in well-being [17].This sample is conducted as a pilot phase of A Longitudinal Study of Egyptian Healthy Aging (AL-SEHA) that adapts the European Survey of Health, Ageing, and Retirement (SHARE) questionnaire.

## 2. Data and methods

### 2.1. Research design

A cross-sectional analytic study was used where all dependent and independent variables are assessed at one and the same point in time.

Our findings are based on questionnaire of A Longitudinal Study of Egyptian Healthy Aging (AL-SEHA) adapting the European Survey of Health, Ageing, and Retirement (SHARE), a multidisciplinary longitudinal survey of noninstitutionalized people aged 50 and up (Borsch-Supan et al., 2005, 2008). Everyone over the age of 50 in the selected households was interviewed. Even if they were under the age of 50, partners of eligible people living in the same household were surveyed. Some questionnaire modules were not distributed to all household respondents.

The so-called “family respondents,” for example, responded to questions about providing child care to grandchildren. These were chosen as the first interviewees in each couple. The order of the interviews within the couple was chosen at random.

### 2.2. Inclusion criteria

Were being able to communicate, and living independently at home, in rented accommodation, in a hostel, or in a retirement home. Those having a gross psychiatric disturbance were excluded. The sample consisted of 299 participants.

A non-probability convenience sampling technique was used in recruiting participants according to the eligibility criteria.

### 2.3. Fieldwork

Investigators from five universities were trained in interviewing and used the prepared SHARE form translated into Arabic to collect the original project data. A sample of eligible elderly people was chosen. The investigators in each governorate contacted potential participants in their area and explained the purpose of the study as well as the data collection procedure. After being informed of their rights, they were invited to participate. Those who agreed to participate were asked to provide a suitable time for the interview. The investigators paid home visits to participants at the appointed time. The data collection form was used to interview them. The data collection procedure took about a month. Each investigator was assigned 15 participants. The collected data were then uploaded to the internet server domain of the American University in Cairo (AUC) Social Research Centre.

### 2.4. Demographic factors

AL-SEHA data set contains detailed data on demographics. Summary statistics of all demographicvariables used in the analyses can be found in Table 1. The demographic factor of interest is family status, which we measure by marital status, the total number of children,children living at home, and grandchildren.Overall governerates 63.54% of the respondents are married, 92.64% have kids, 47.49% have grandchildren, and 77.92% are living with their kids in the same house or the same building.

**Table 1.** Sample demographics

|  | N | % |
| --- | --- | --- |
| **Gender** |  |  |
| Male | 128 | 42.81 |
| Female | 171 | 57.19 |
| **Education** |  |  |
| No education | 48 | 16.05 |
| Primary education | 31 | 10.37 |
| prep education | 18 | 6.02 |
| High school | 48 | 16.05 |
| Medium high | 18 | 6.02 |
| Bachelor | 112 | 37.46 |
| Post grad | 24 | 8.03 |
| **Age** |  |  |
| 50-60 | 151 | 50.5 |
| 60-70 | 88 | 29.4<br>3 |
| 70-80 | 50 | 16.7<br>2 |
| 80-90 | 8 | 2.68 |
| above 90 | 2 | 0.67 |
| **Marital Status** |  |  |
| Not married | 5 | 1.67 |
| Engaged | 0 | 0 |
| Married | 190 | 63.5<br>5 |
| Separated | 3 | 1 |
| Divorced | 13 | 4.35 |
| Widow | 88 | 29.4<br>3 |
| **Parental status** |  |  |
| No kids | 22 | 7.36 |
| Kids >=1 | 277 | 92.64 |
| **Grandparenting status** |  |  |
| No grandkids | 87 | 29.1 |
| Grandkids>=1 | 142 | 47.49 |

### 2.5. Family networks

Respondents are asked to answer questions about their family support network. Including their marital status, having children and grandchildren, frequency of contact and proximity with their family members.

Each respondent’s marital status is classified as, (1) never married, (2) married and living together with the spouse,, (3) divorced, and (4) widowed. For the regression analysis, we create the dummy variables married (one if the respondent is married or in a registered partnership), divorced (one if the respondent is divorced or living separated from spouse), and widowed (one if the respondent is widowed).

We define the two-category measure of children as no children, or having children. We define two category measure of having grandchildren as have grandchildren or not having grandchildren..

For contact frequency, the respondent is asked how much contact he had with each person in his social support network in the previous 12 months. (1) daily, (2) several times a week, (3) approximately once a week, (4) approximately every two weeks, (5) approximately once a month, (6) less than once a month, and (7) never. After clearing the outliers, We recode the measure so that it ranges from 1 (daily or weekly) 2 (twice a week) and 3 (once a month or less).

For proximity, similar measures are constructed. The respondent is asked how far the individual lives and proximity is classified as (8) other country, (7) more than 500km, (6) 100km to 500km, (5) from 25 km 100km, (4) 15km to 25km, (3) 1km to 15km, and (2) less than 1km, (1) living in the same building, (0) same house/apartment. After clearing outliers, we recoded the measure that the respondent have at least one child who lives in (1) the same house or same building, (2) between 1 km to 15 km, and (3) more than 15 km.

### 2.6. Cognitive function

The data were extracted from the SPSS (Version 25) dataset for statistical analysis. Bivariate analyses included chi-square tests. As appropriate, data were compared using independent t-tests.

### 2.7. Dependant variable

A composite measure based on three cognitive tests of immediate recall, delayed recall, and fluency was used to assess cognitive performance (Adam, Bonsang, Grotz, & Perelman, 2013).

The immediate and delayed recall tasks are episodic memory tasks that assess short-term verbal learning, memory, and information retention (Cheke & Clayton, 2013).

The interviewer reads aloud a list of ten words and asks the respondent to recall as many as he or she can during the immediate memory task. The respondent is asked to repeat these words 5-10 minutes later in the delayed recall task. Each of these tests has a score range of 0 to 10. Respondents were asked to rate their verbal fluency, which is a measure of executive function and language ability (Henry, Crawford, & Phillips, 2004). name as many animals as they can in one minute. The composite measure of cognition was constructed by averaging the standardized scores of these three indicators to range between 0 and 1, where between 0 and 0.5 is considered impaired and between 0.5 and 1 is considered unimpaired.

### 2.8. Covariates and moderators

A variety of factors influence cognitive performance in later life. Age, education, gender, marital status are all socioeconomic and health variables to consider (Litwin & Stoeckel, 2015).

As a result, we included these variables as covariates in our analyses.

Gender was used as a dummy variable (1) Men, (2) Women and age was used as a dummy variable as (1) between 50 and 60, (2) between 60 and 70, (3) more than 70.

Years of education were used as a dummy variable as (1) no education, (2) basic to intermediate, (3) university or postgraduate studies to measure education.

### 2.9. Analyses of data

To begin, descriptive statistics were used to compute The means and standard deviations of continuous variables, as well as the percentages and frequencies of categorical variables.

Bivariate analyses were performed in the second stage to examine the relationship between cognitive health and the study variables, using independent t-tests for factors.

### 2.10. Cognitive performance

The researchers assessed cognitive functioning in two domains derived from three cognitive tasks: verbal fluency, immediate word recall, and delayed word recall. These three measures indexed executive function and memory constructs.

Executive power. A category fluency task was used to assess executive function, in which participants were asked to name as many animals correctly as possible in one minute.

Participants must devise a strategy for recalling category exemplars, which is an assessment of executive functioning. It has been widely used as a component of neuropsychological batteries to differentiate between healthy age-related memory change and clinically significant impairments (Haugrud, Crossley, & Vrbancic, 2011). Memory function was evaluated using the Ten-Word Delayed Recall Test’s immediate and delayed word recall. Ten common words were presented to participants, who were then asked to recall them immediately and again five minutes later. The memory measure was created by averaging the two scores. This assessment was built on similar computerized word recall tasks that have been widely used to assess both immediate and delayed memory performance (Green, Montijo, & Brockhaus, 2011; Hoskins, Binder, Chaytor, Williamson, & Drane, 2010).

## 3. Results

Al-SEHA questionnaire contained detailed demographic data. We focused in our study on the family characteristics analyzed in Table 1. The family status is measured by the marital status and total number of children. In addition, personal characteristics were also measured, including gender, age, and education status to be used as controls for the analysis. Our dataset included 299 respondents, 128 men (42.81%) and 171 women (57.19%). Ages ranged from 50 to more than 90, with 50.5% of ages 50-60, 29.43% of ages 60-70, 16.72% of ages 70-80, 2.68% of ages 80-90 and only 2 respondents (0.67%) aged more than 90 years. In addition, the majority of respondents (37.46%) hold bachelor degrees, while only 16.08% received no education. Yet, 10.37%, 6.02%, 16.02% of them received primary education, preparatory education and high school degree respectively. Only 24 respondents (8.03%) holded post graduate degrees. Table 1. Summary statistics of demographic variables Most of the respondents reported that they got married before. Only 5 cases (1.67%) never got married. The marital status of each respondent was classified into categories: (1.67%) not married, (0%) engaged,(63.55%) married, (1%) separated, (4.35%) divorced, (29.43%) widow. (92.6%) reported having more than one child and (47.49%) reported having more than one grandchild.

### 3.1. Cognitive functions

In the recall tests, which measured working memory, the interviewer first read a list of ten common words to the respondent and then asked the respondent to recall as many words from the list as possible in any order (immediate recall). Recall was allowed for up to one minute. The test was repeated at the end of the cognitive function module, but the words were not read aloud again (delayed recall).

The range was 1 to 19, with an average of about 6 animals named within 1 minutes (for men: M=6.3, SD=4.07; for women: M=6.13, SD=3.92

For the immediate recall, the mean number of words is about 5 words with SD=2.17 (For men: M=5.8, SD=2.19; for women: M=5.47, SD=2.14). 71.5% of the respondents were able to recall more than five words immediately after hearing them.

For the delayed recall, the mean number of words is about 4 words with SD=2.21 (For men: M=4.67, SD= 2.23; for women: M= 4.25, SD=2.16). 46.1% of the respondents were able to recall more than 5 words after some time. Figure 1. represents results of overall cognitive functions per gender and Figure 2. Represents visualization of overall cogntive functions per age and gender.

**Figure 1.**
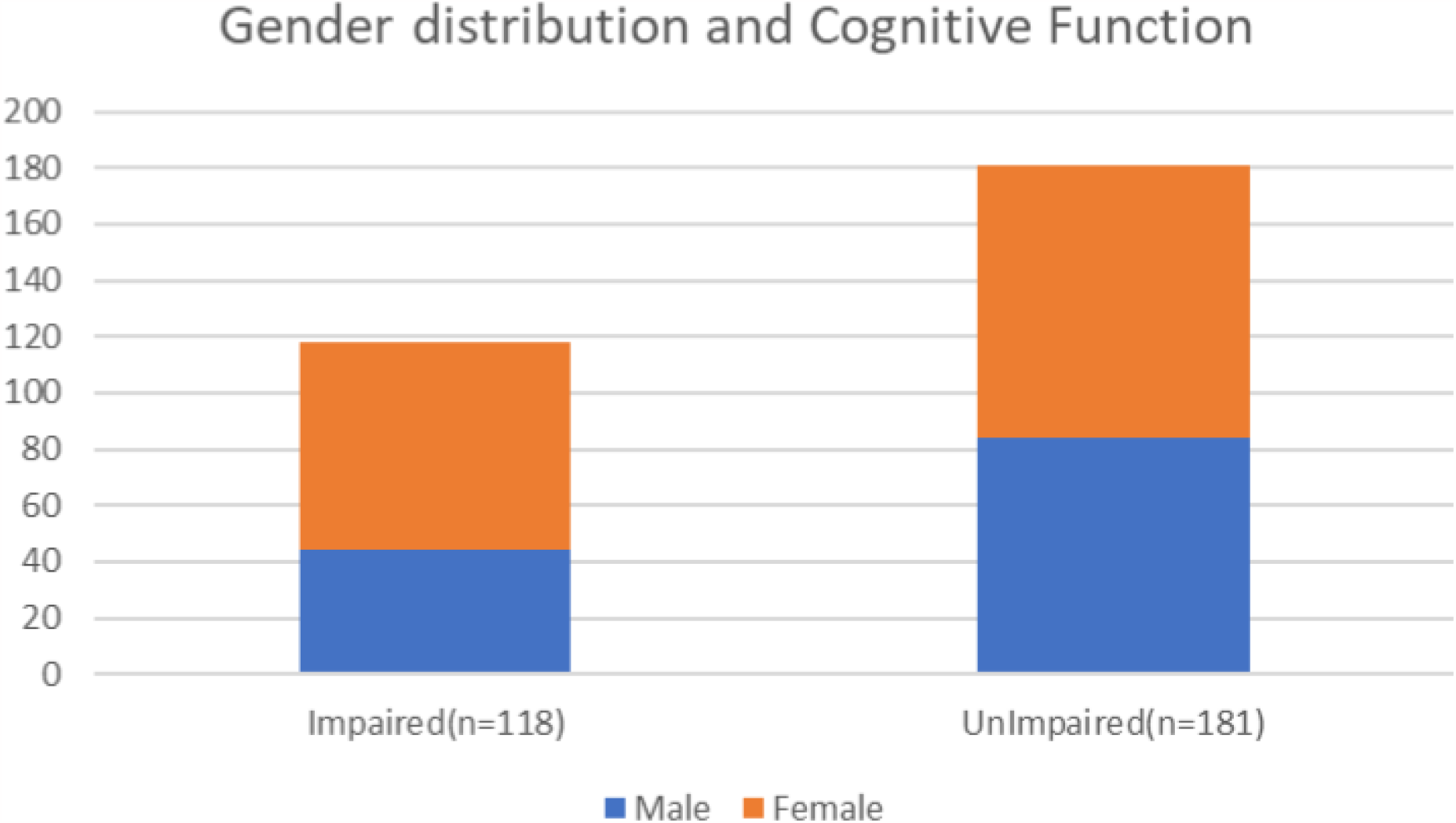
Cognitive functions according to gender

**Figure 2.**
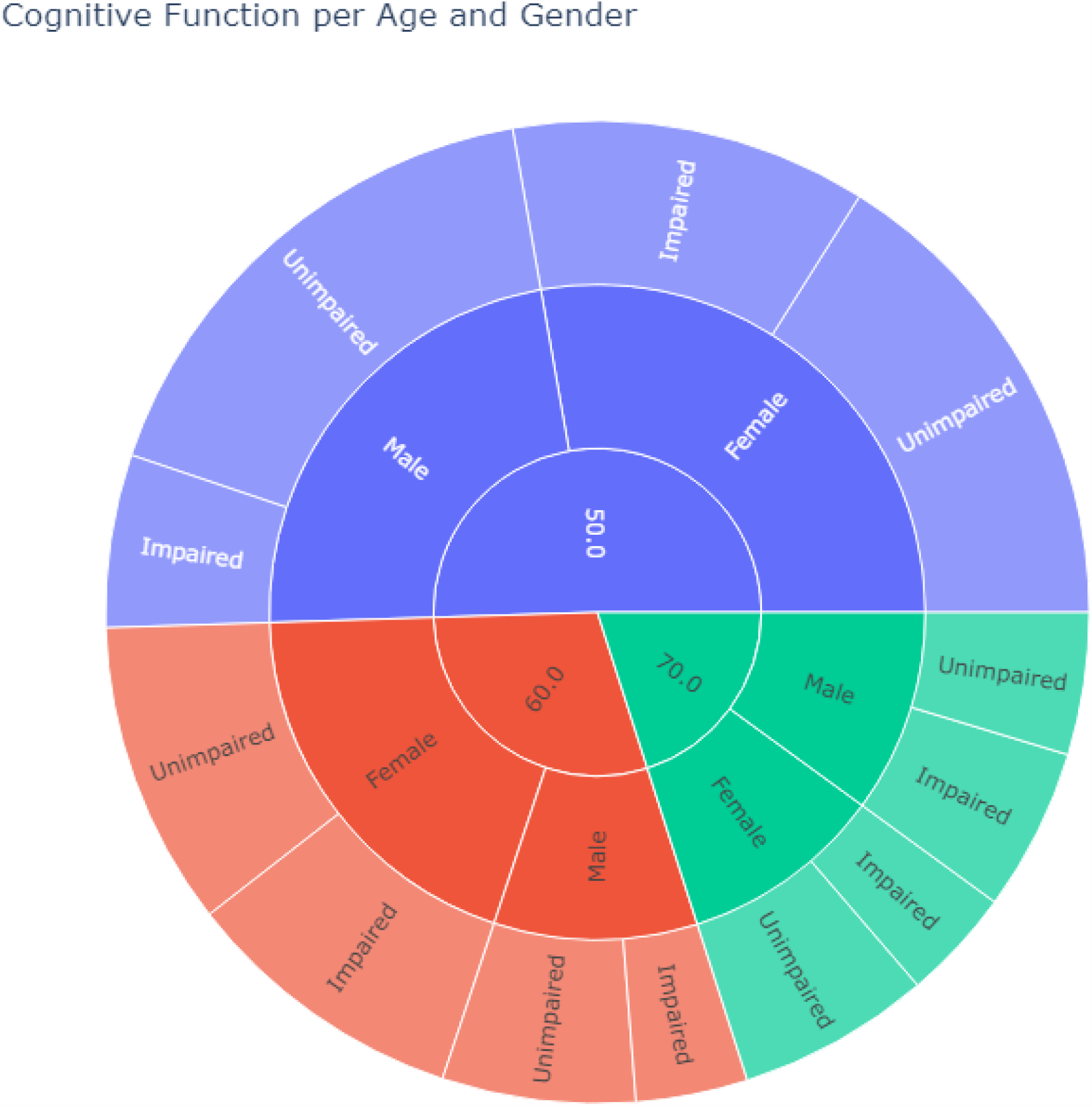
Visualization of cognitive functions per age and gender

Table 2. Measures cognitive functions against age, gender and education. Where improved cogntive functions are positively correlated with education as represented in Figure 3., with a p value of **0.0001**. Compared to 10.5% of those who received no education.

**Table 2.**
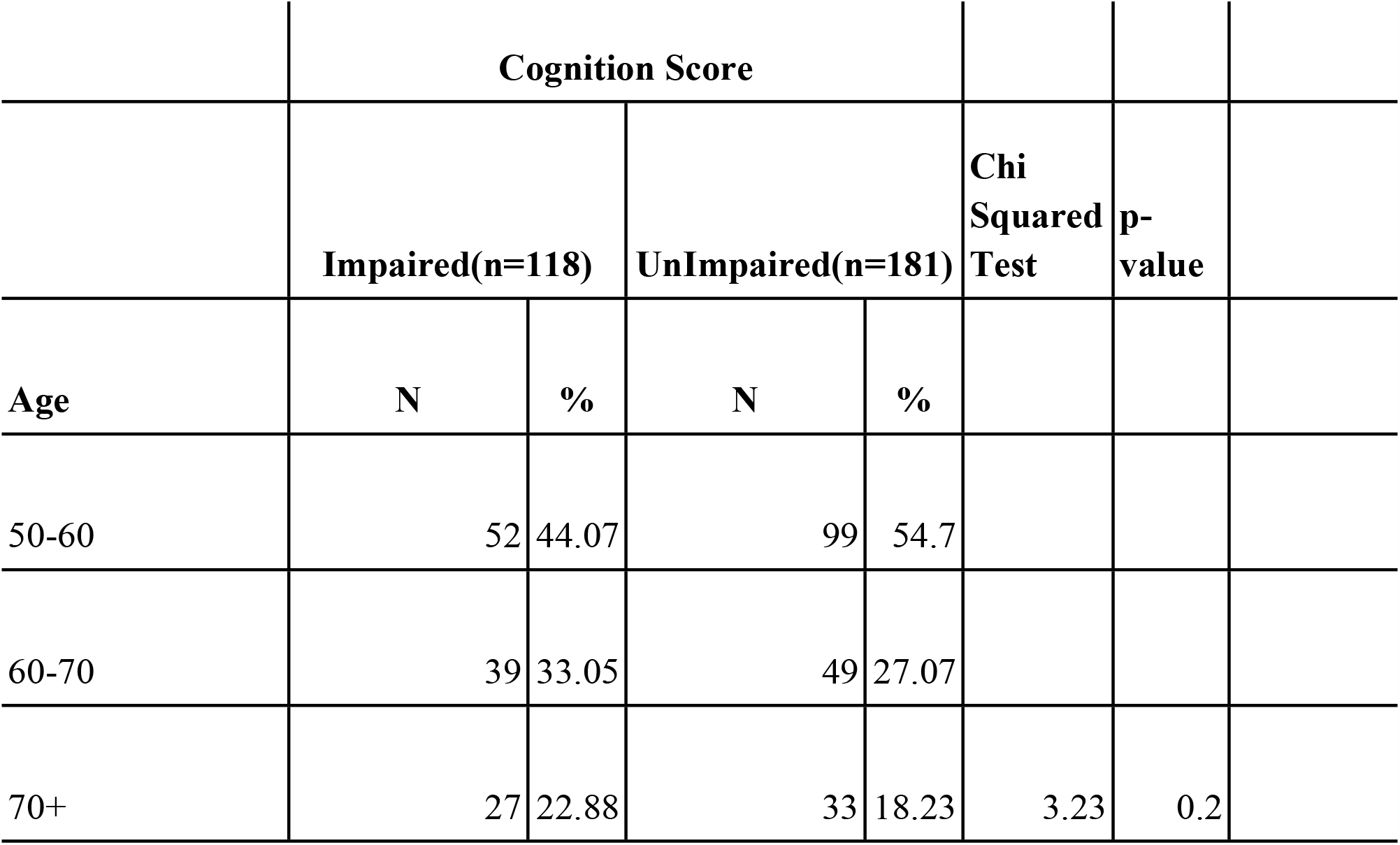
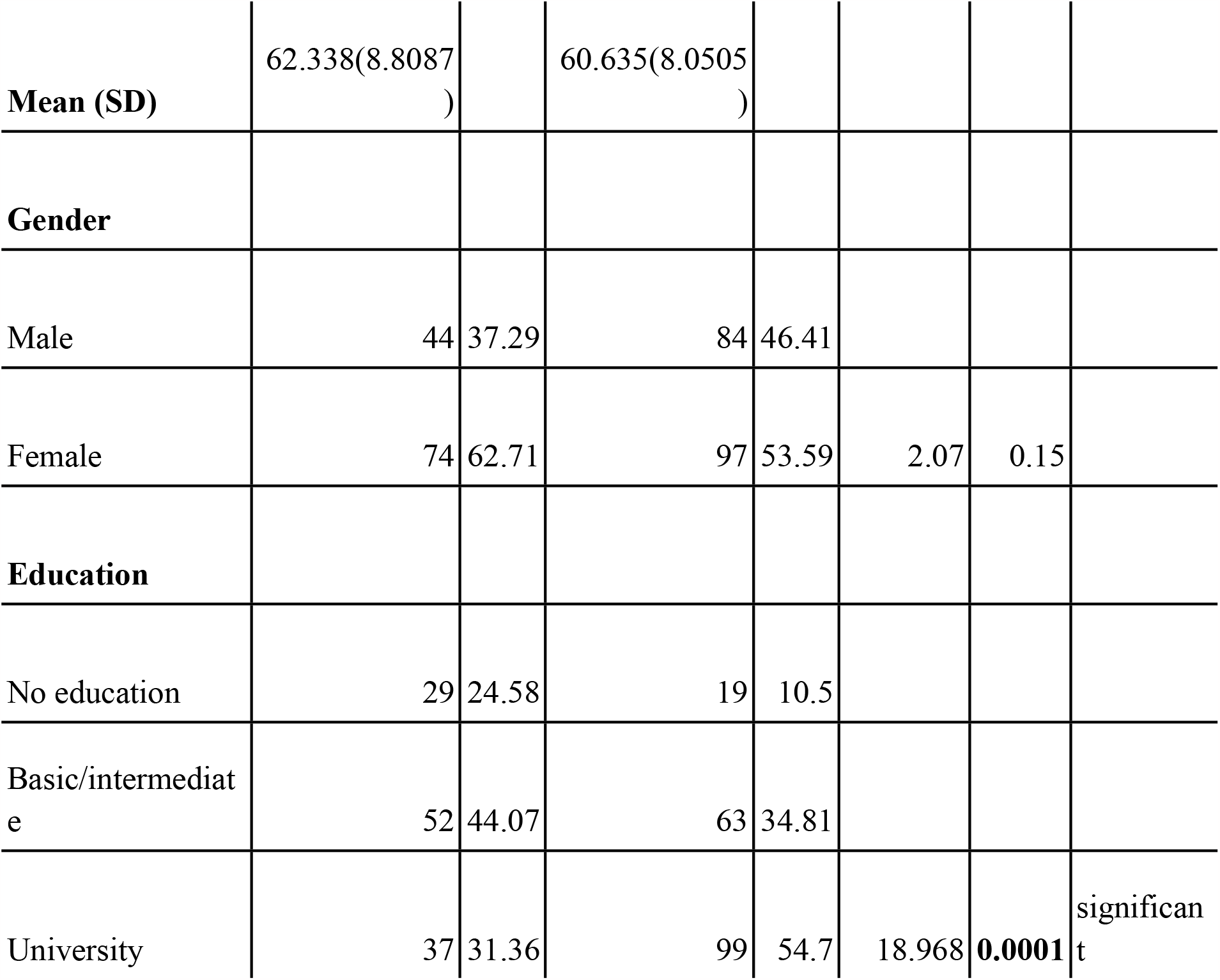
Cognitive functions with age, gender and education level

**Figure 3.**
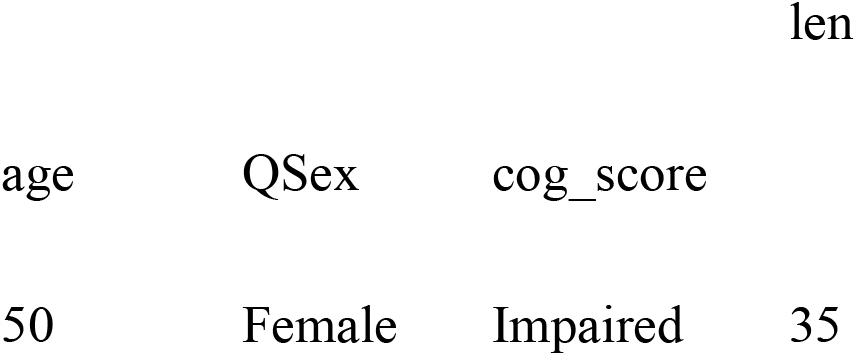
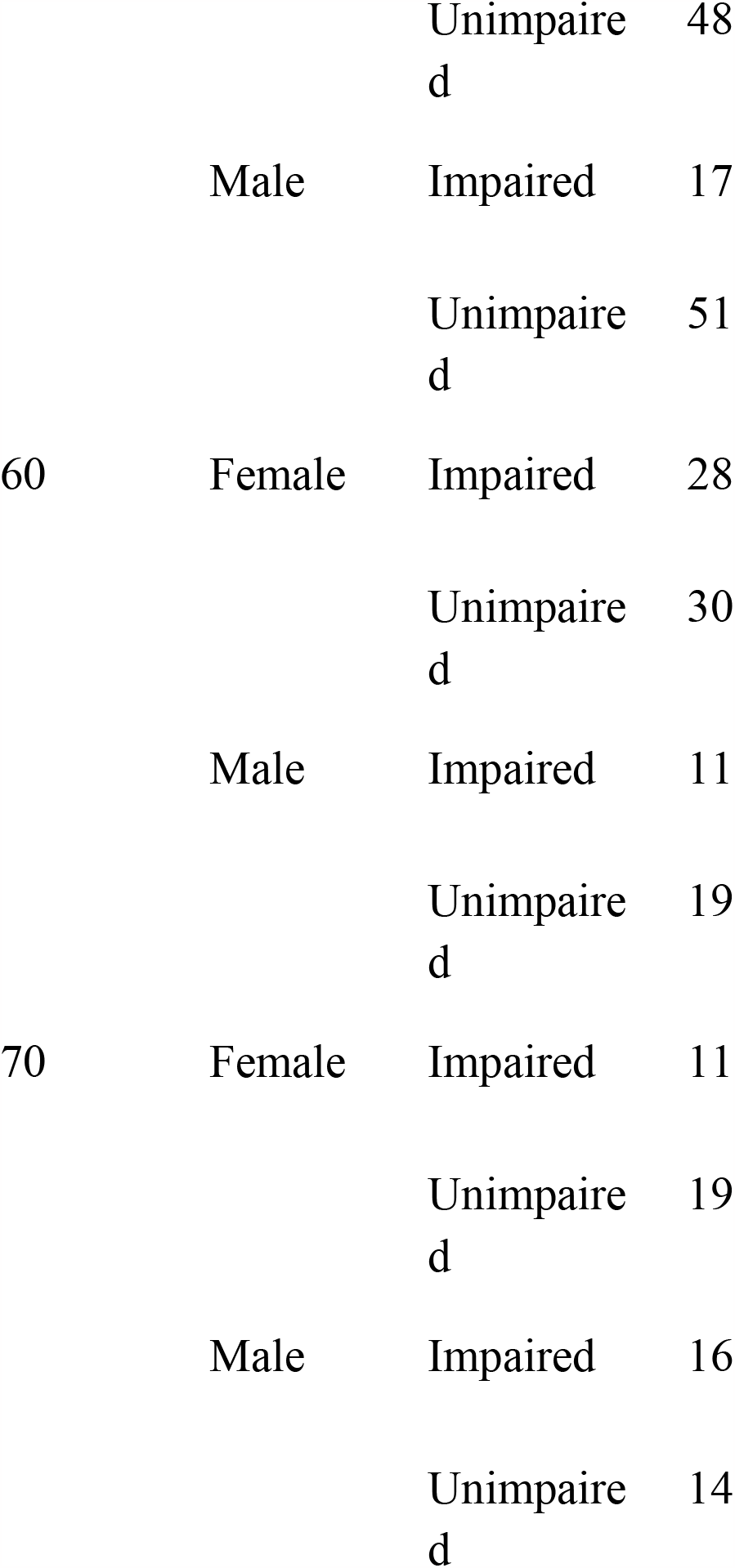
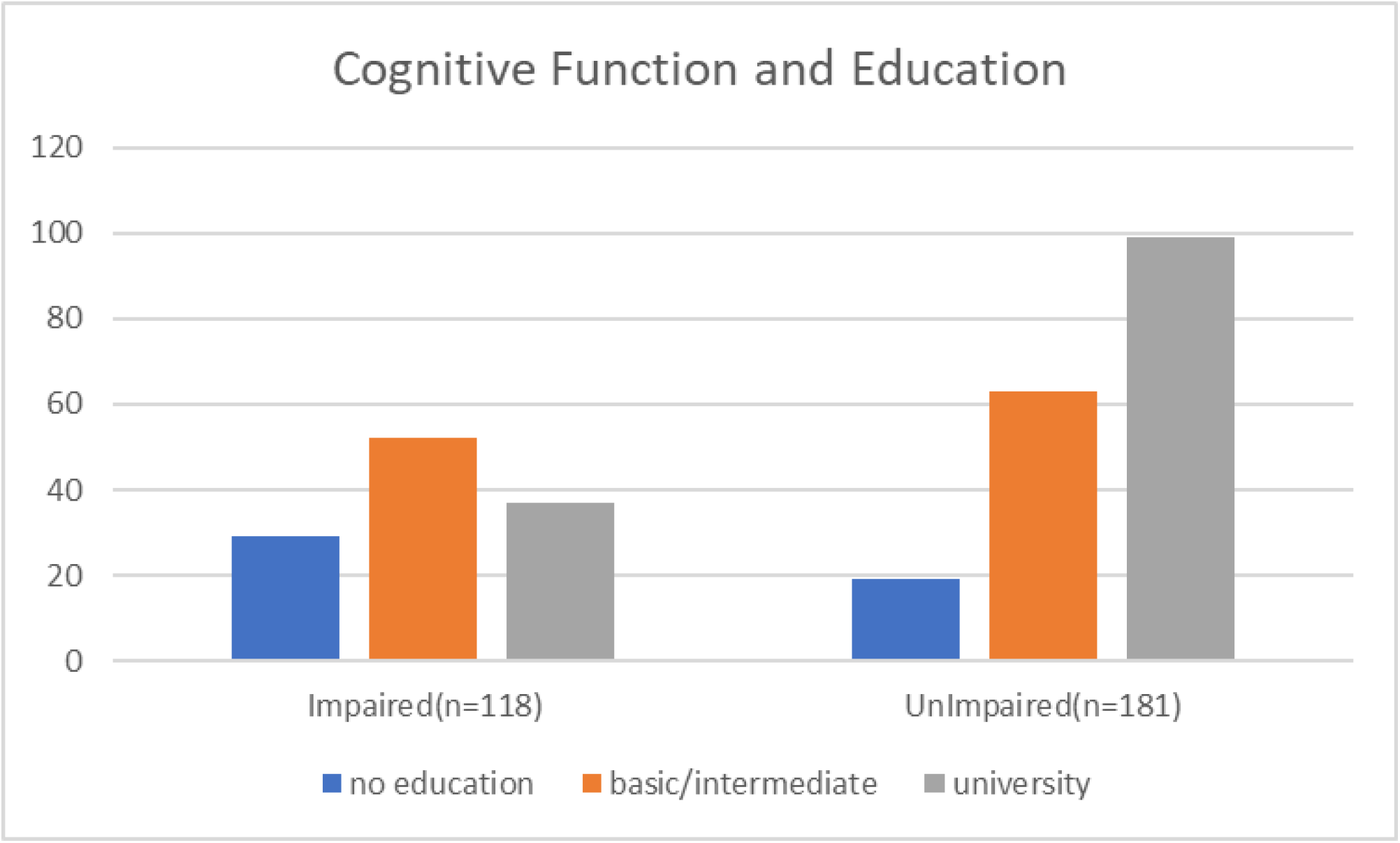
Relationship between cognitive functions and level of education

As for marital status, 53.59% of females were impaired compared to 46.41% of males as represented in Figure 4.. The age group between 50-60 showed the highest improved cognitive functions with 54.7% compared to 18.23% at the age group of 70+ with a p value of **0.0378**.

**Figure 4.**
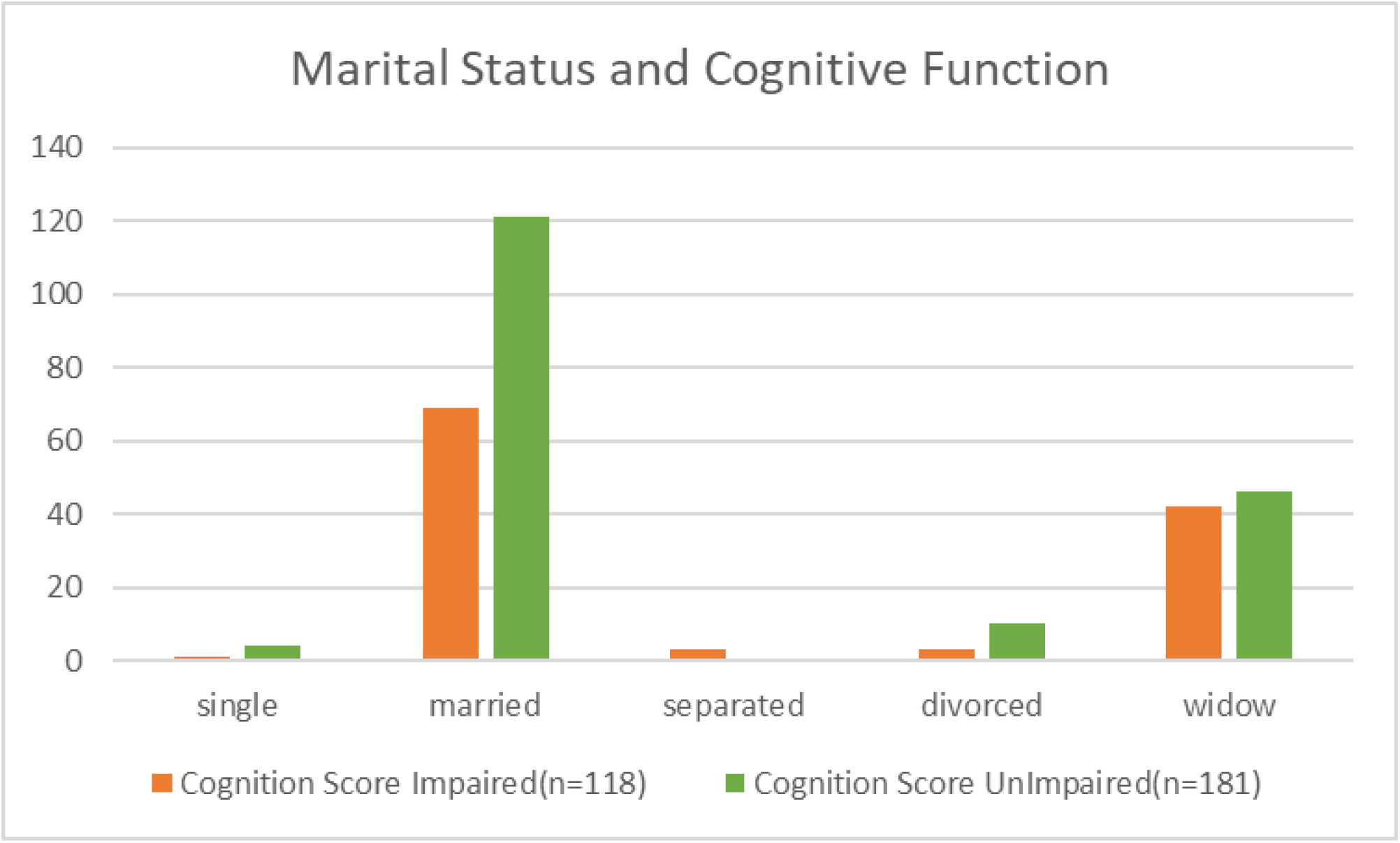
Relationship between cognitive functions and marital status

Understanding the effect of marital status, proximity, contact frequency, care of grandchildren and getting help from family members is reported in Table 3.

**Table 3.**
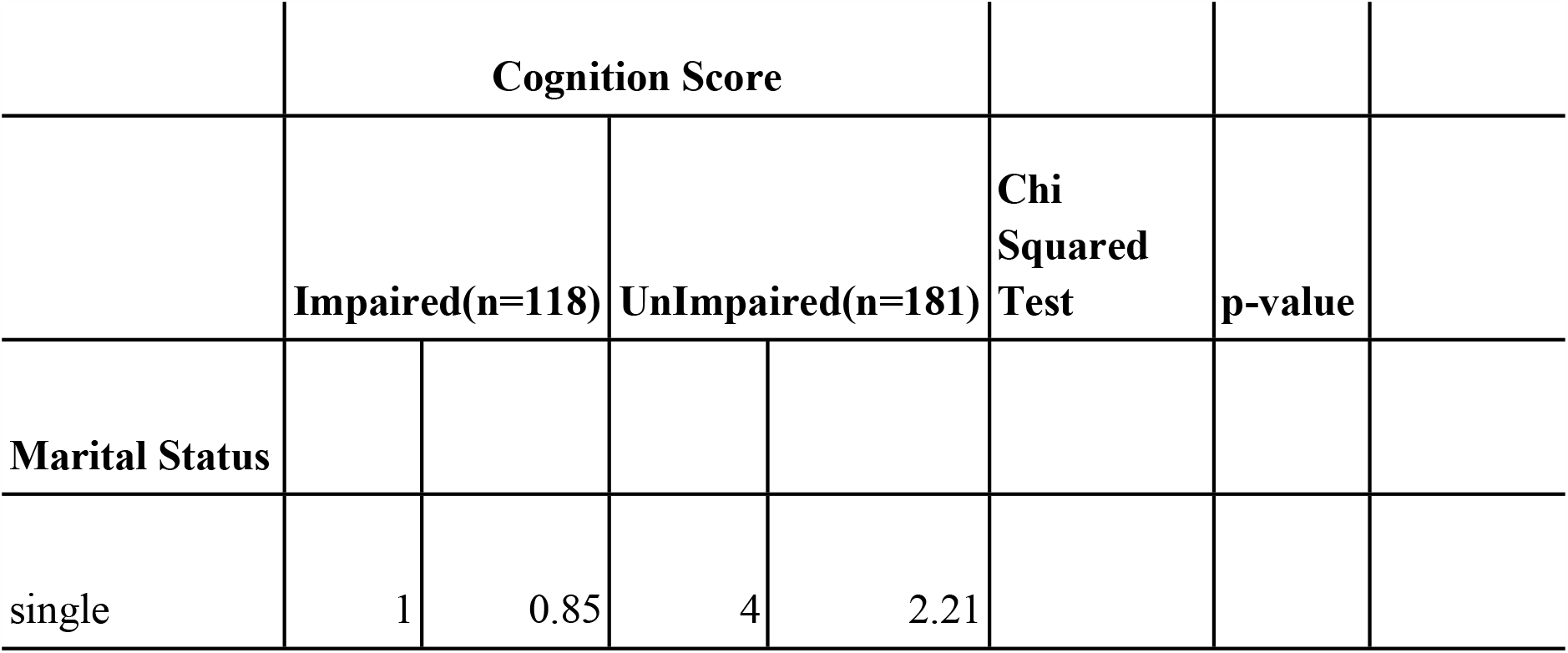
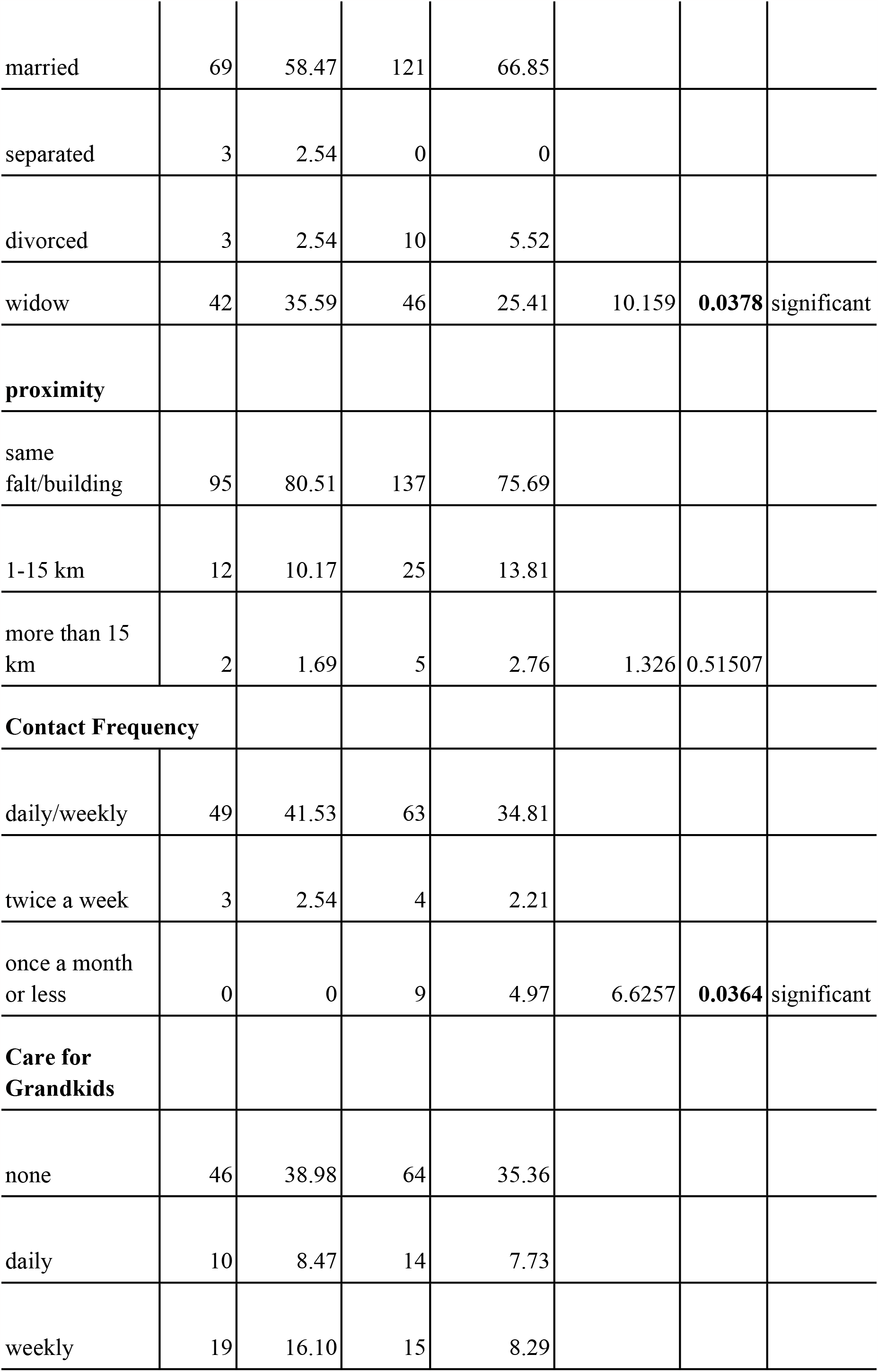
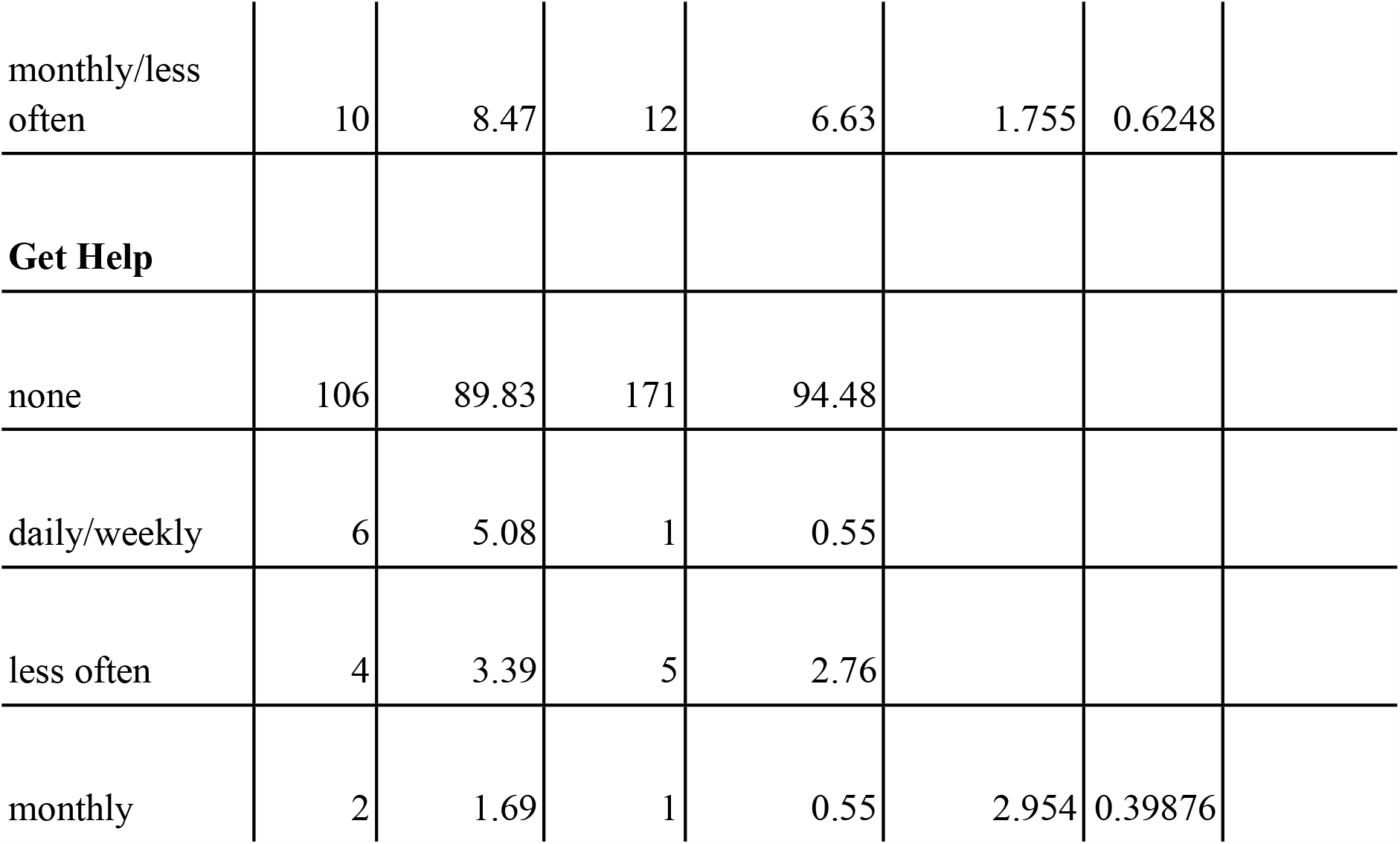
Relationship between cognitive functions and marital status, proximity, contact frequency, care for grandkids and receiving help from others

|  | Cognition Score |  |  |  |  |  |
| --- | --- | --- | --- | --- | --- | --- |
|  | Impaired(n=118) |  | UnImpaired(n=181) |  | Chi Squared Test | p-value |
| **Marital Status** |  |  |  |  |  |  |
| single | 1 | 0.85 | 4 | 2.21 |  |  |

|  |  |  |  |  |  |  |  |
| --- | --- | --- | --- | --- | --- | --- | --- |
| married | 69 | 58.47 | 121 | 66.85 |  |  |  |
| separated | 3 | 2.54 | 0 | 0 |  |  |  |
| divorced | 3 | 2.54 | 10 | 5.52 |  |  |  |
| widow | 42 | 35.59 | 46 | 25.41 | 10.159 | **0.0378** | significant |
| **proximity** |  |  |  |  |  |  |  |
| same<br>falt/building | 95 | 80.51 | 137 | 75.69 |  |  |  |
| 1-15 km | 12 | 10.17 | 25 | 13.81 |  |  |  |
| more than 15<br>km | 2 | 1.69 | 5 | 2.76 | 1.326 | 0.51507 |  |
| **Contact Frequency** |  |  |  |  |  |  |  |
| daily/weekly | 49 | 41.53 | 63 | 34.81 |  |  |  |
| twice a week | 3 | 2.54 | 4 | 2.21 |  |  |  |
| once a month<br>or less | 0 | 0 | 9 | 4.97 | 6.6257 | **0.0364** | significant |
| **Care for**<br>**Grandkids** |  |  |  |  |  |  |  |
| none | 46 | 38.98 | 64 | 35.36 |  |  |  |
| daily | 10 | 8.47 | 14 | 7.73 |  |  |  |
| weekly | 19 | 16.10 | 15 | 8.29 |  |  |  |
| monthly/less often | 10 | 8.47 | 12 | 6.63 | 1.755 | 0.6248 |
| **Get Help** |  |  |  |  |  |  |
| none | 106 | 89.83 | 171 | 94.48 |  |  |
| daily/weekly | 6 | 5.08 | 1 | 0.55 |  |  |
| less often | 4 | 3.39 | 5 | 2.76 |  |  |
| monthly | 2 | 1.69 | 1 | 0.55 | 2.954 | 0.39876 |

Out of n=181 respondents, married partiticapnts (66.85%) identified as unimpared. As for proximity to children, 75.67% of unimpared respondents lived in the same flat/building with their children.

34.81% of unimpaired respondents, reported to have a daily/weekly contact with their children, with significance of p value **0.0364** as represented in Figure 5. While not having any grandchildren and not receiving any help from family members, showed highest percentage of unimpared respondents with 35.36%, 94.3% respectively.

**Figure 5.**
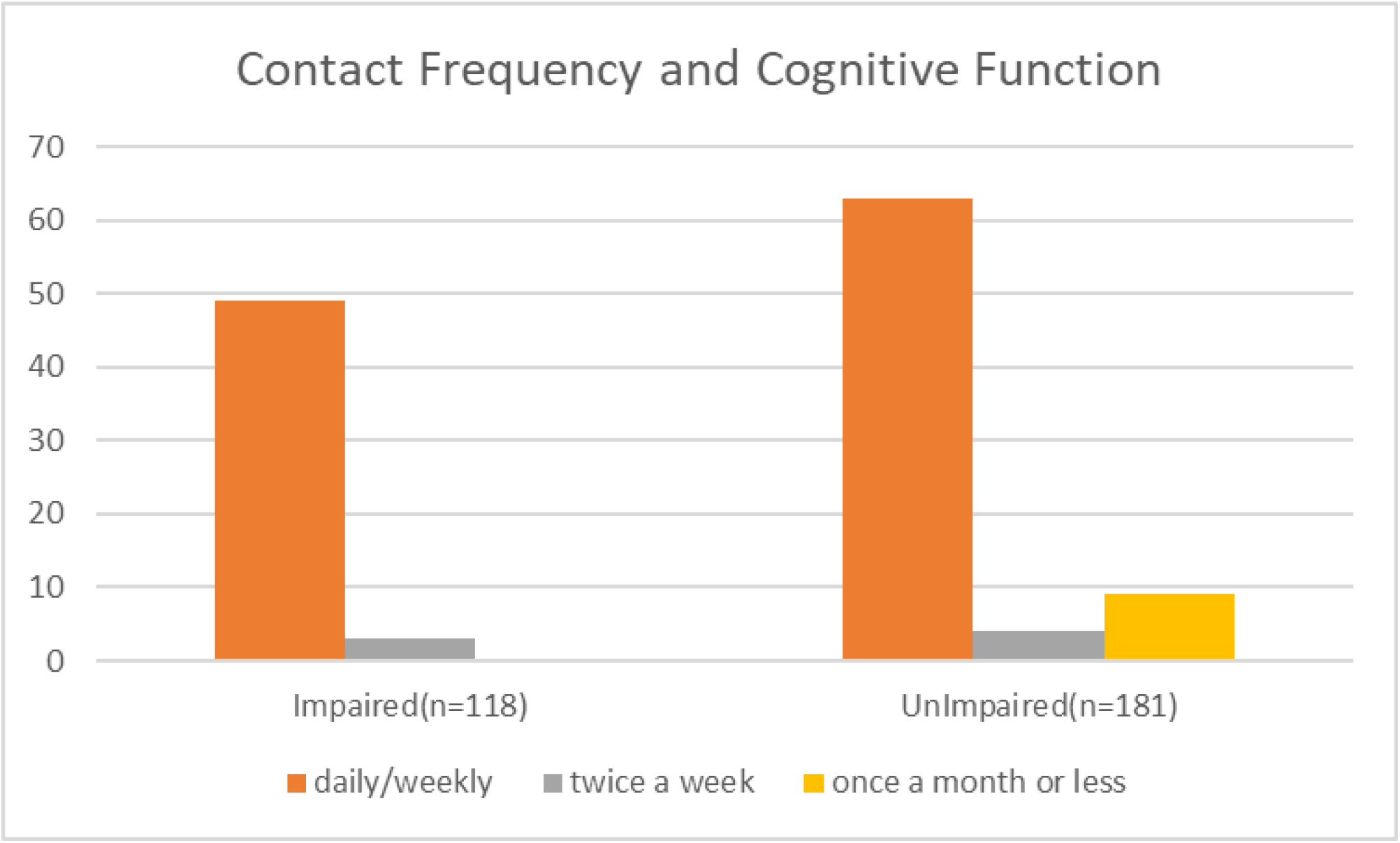
Relationship between cognitive functions and contact frequency with children

## 4. Discussion

This study aims at understanding the effect of family network and support on cognitive functions at the Egyptian population, using adopted SHARE questionnaire by AL-SEHA study.

Measuring cogntive functions was done through three tests according to the SHARE questionnaire. Verbal fluency, immediate and delayed recall. Although knowledge is important in all cognitive tests (Hertzog, 2008), the delayed recall measure of cognitive performance relies primarily on recently stored information. Furthermore, both recall measures require the acquisition of new information (i.e., a list of objects to be repeated)

The verbal fluency test captured the crystallized/knowledge components of cognition in our study, representing products of past processing.

We acknowledge that the verbal fluency test we used may have been influenced by processing speed, and that better measures of (pure) crystallised abilities could be used.

As for our results, we measured various variables and their impact on cogntive functions.

There was a statistically significant relationship between education, marital status and contact frequency with children and positive cognitive functions. These findings support our hypothesis that social support in Egyptian older adults is a predictor of cognitive function.

We believe that interventions or activities that increase social support would benefit cognitive functions in the elderly.

Analyzing various variables as marital status, education level with cognitive functions as well as family support represented as proximity to children, contact frequency, caring for grandchildren and getting help from family memers.

As for level of education, our results showed a positive correlation between level of education and cognitive functions. Out of different levels of education ranging from no education to university/post graduate degree, possessing a university/post graduate degree reported the highest level of cognitive functions with strong significance and p value of <**0.0001**.

Literature showed mixed reports on the effect of having a partner on cogntive functions.Our results were sided with the literature reporting improved cogntive functions, as it showed a significance when correlating marital status with cogntive functions as reported in table 2., those who are married reported higher levels of cognitive functions with p value= **0.0378**.

Furthermore, the ‘use it or lose it’ theory holds that contact frequency has a direct impact on cognitive health in old age. This viewpoint holds that frequent interaction with others provides ‘cognitive exercise,’ stimulating the mind and maintaining cognitive functions. Such encounters can also improve cognitive reserve, allowing people to tolerate brain pathology while exhibiting no obvious behavioural symptoms. We studied The impact of contact frequency with children on cognitive functions, where it aligned with the literature where a positive effect on participants’ cognitive functions was reported. With significance and p value =**0.0364**.

Taken together, our findings suggest that family support may be important for old-age cognitive functions. Spouses and children are frequently the foundation of long-lasting social networks that can provide social support to the elderly.

More research is needed to determine which factors cause the positive effects found here, such as comparing well-being and mental health before and after the formation of partnerships or social networks in longitudinal data.

Among those considered, the verbal fluency test in SHARE has the largest knowledge component and primarily represents products of previous processing.

## 5. Conclusion

In conclusion, this research contributes to our understanding of the impact of family network and support and cognitive function in the older Egyptian population. Our findings can be a base to add on to the literature.

Demonstrating the effect of different variables such as marital status, level of education, contact frequency with children, taking care of grandchildren, receiving help from others, and proximity to children on cognitive function. Our results showed a positive correlation between marital status and being married to better cognitive functions. People with a university degree or higher, possesses unimpaired cognitive functions compared to others.

Increased Contact frequency showed a positive correlation to cognitive functions in our sample. This study paves the way for longitudinal studies to better understand the correlation between these variables and cognitive functions in the Egyptian population over time, and the development of culturally sensitive and more effective forms of interventions to slow age-related cognitive decline and improve older people’s quality of life.

## Data Availability

The original contributions presented in the study are included in the article/supplementary material, further inquiries can be directed to the corresponding author/s.

## Author contribution statement

**SAM, NG**,**SH:** Drafted the first version, conceptualized the idea, conducted data collection and analysis.

**MS**: Funding, conceptualization, final revision

## Ethics statements

This study was reviewed and approved by IRB of the American University in Cairo (IRB-AUC). The participants provided their written informed consent to participate in this study.

## Inclusion of identifiable human data

No potentially identifiable human images or data is presented in this study.

## Data availability statement

Generated Statement: The original contributions presented in the study are included in the article/supplementary material, further inquiries can be directed to the corresponding author/s.

## STATEMENTS

### Data availability statement

All data are available upon request from the corresponding author.

### Ethics approval statement

The current work has been approved by the IRB of the American University in Cairo. IRBAUC_ Case# 2021-2022-029

### Patient consent statement

All recruited subjects signed an informed consent form to join the study.

